# Clinical Value Of CT-Derived Simulations Of Transcatheter-Aortic-Valve-Replacement In Challenging Anatomies The PRECISE-TAVR Trial

**DOI:** 10.1101/2023.02.10.23285640

**Authors:** Thijmen W Hokken, Hendrik Wienemann, James Dargan, Dirk-Jan van Ginkel, Cameron Dowling, Axel Unbehaun, Johan Bosmans, Andreas Bader-Wolfe, Robert Gooley, Martin Swaans, Stephen J. Brecker, Matti Adam, Nicolas M. Van Mieghem

**Author notes:** **Address of correspondence:** Nicolas M. Van Mieghem, MD, PhD, Department of Interventional Cardiology, Thoraxcenter, ErasmusMC, Office Nt 645, Dr Molewaterplein 40 3015 GD Rotterdam, The Netherlands.

## Abstract

**Background:** Pre-procedural computed tomography planning improves procedural safety and efficacy of transcatheter aortic valve replacement(TAVR). However, contemporary imaging modalities do not account for device-host interactions. This study evaluates the value of pre-procedural computer simulation with FEops HEARTguide™ on overall device success in patients with challenging anatomies undergoing TAVR with a contemporary self-expanding supra-annular transcatheter heart valve.

**Methods:** This prospective multicenter observational study included patients with a challenging anatomy defined as bicuspid aortic valve, small annulus or severely calcified aortic valve. We compared the heart team’s transcatheter heart valve(THV) planning decision based on 1) conventional multislice computed tomography(MSCT) and 2) MSCT imaging with FEops HEARTguide™ simulations. Clinical outcomes and THV performance were followed up to 30 days.

**Results:** A total of 77 patients were included(Median age 79.9 years (IQR 74.2-83.8), 42% male). In 35% of the patients, pre-procedural planning changed after FEops HEARTguide™ simulations(change in valve size selection(12%) or target implantation height(23%)). A new permanent pacemaker implantation(PPI) was implanted in 13% and >trace paravalvular leakage (PVL) occurred in 28.5%. The contact pressure index(i.e. simulation output indicating the risk of conduction abnormalities) was significantly higher in patients with a new PPI, compared to those without(16.0%(25^th^-75^th^ percentile 12.0-21.0) vs. 3.5%(25th-75th percentile 0−11.3), p<0.01) The predicted PVL was 5.7mL/s(25^th^-75^th^ percentile 1.3-11.1) in patients with none-trace PVL, 12.7(25th-75th percentile 5.5-19.1) in mild PVL and 17.7(25^th^-75^th^ percentile 3.6-19.4) in moderate PVL(p=0.04).

**Conclusion:** FEops HEARTguide™ simulations may provide enhanced insights in the risk for PVL or PPI after TAVR with a self-expanding supra-annular THV in complex anatomies.

## Introduction

Transcatheter Aortic Valve Replacement (TAVR) is recommended for elderly patients with symptomatic severe aortic stenosis (AS) across the operative risk spectrum.(1,2) Electrocardiogram(ECG)-gated, contrast-enhanced multi-slice computed tomography (MSCT) is the preferred imaging modality for obtaining detailed information on aortic valve morphology, dimensions, calcium burden, membranous septum length, left ventricular outflow tract (LVOT) and transcatheter heart valve sizing.(3,4) The CoreValve/Evolut platform is a widely used self-expandig supra-annular transcatheter valve that compared favorably to SAVR in a series of randomized controlled trials with superior hemodynamic valve performance but with more paravalvular leakage and a higher need for new pacemaker implantation.(5-7)

Heavily calcified tricuspid, bicuspid and small aortic valves may pose specific challenges from a TAVR perspective and are associated with paravalvular leakage (PVL), conduction abnormalities and aortic root injury. (8-10).

Insights in device-host interactions may help to understand and predict such TAVR related complications. FEops HEARTguide™ is a CE marked software package, also regulatory approved in Canada and Australia, for patient-specific simulations for structural heart interventions, which can support to determine the risk for conduction abnormalities and PVL post-TAVR by virtually implanting a transcatheter heart valve in a 3D anatomical computer model. The computer simulations has been validated in tricuspid and bicuspid anatomies.(11-14)

The goal of this prospective multi-center observational study (PRECISE-TAVR) is to evaluate the effect of FEops HEARTguide™ on THV sizing and implantation strategy in severe AS patients with challenging aortic anatomy in order to predict risk of PVL and conduction abnormalities following TAVR with the Evolut Pro(+).

## Methods

### Study Population

The PRECISE-TAVR trial is a prospective multicenter observational study, including patients with a challenging anatomy, eligible for an Evolut Pro valve (Medtronic, Minneapolis, MN). A challenging anatomy was defined as 1) a bicuspid aortic valve, 2) a heavily calcified tricuspid valve (with Agatston score >3000 for men and >1600 for women) or 3) a small aortic valve (mean annular diameter <20mm). Formal exclusion criteria were presence of a permanent pacemaker prior to TAVR, a failing surgical bio-prosthesis or suboptimal MSCT imaging quality precluding accurate computational modelling. The study was conducted in accordance with the declaration of Helsinki and did not fall under the scope of the Medical Research Involving Human Subjects Act per Institutional Review Boards’ review (MEC-2020-0486).

### Study Procedure

First, multi-disciplinary heart teams identified patients with challenging aortic anatomies who were selected for TAVR with the Evolut Pro (+) THV based on MSCT analyses per local standard. THV sizing and implantation strategy were documented. Second, patient-specific computer simulations of device implantation were performed and the derived contact pressure and PVL were obtained. Simulations were then shared with the local heart teams. THV sizing and implantation strategies could be changed accordingly per local heart team’s discretion.

A dedicated prospective database captured relevant patient demographics, medical history and comorbidities, ECG, Transthoracic Echocardiography(TTE) and MSCT findings including THV sizing and implantation strategies before and after FEops HEARTguide™ simulations, procedural and clinical follow up data.

### Computer simulations

MSCT imaging studies were transmitted to FEops (Gent, Belgium) for HEARTguide computer simulations of device implantation. A detailed description of the computer simulations has been described earlier.(14) In brief, a 3-dimensional, patient-specific aortic root was reconstructed from the pre-procedural ECG-gated contrast-enhanced CT-scan with finite element models. For each patient, Evolut TAVR simulations were performed for the 2 most appropriate available device sizes and at an implantation depth < 3mm (high implantation) and 5mm (medium implantation). The THV properties of the models were assessed from micro-CT images and optical microscopy measurements, as well as in-vitro radial compression tests at body temperature.

Computer simulations have been already used to predict the risk of conduction abnormalities and PVL post-TAVR. For the prediction of conduction abnormalities, the contact pressure exerted on the region nearby the membranous septum is extracted from the simulation. More in detail, the region of interest is defined as extending from the inferior border of the membranous septum to 15mm below the annulus. This is the area where the HIS-bundle surfaces in the LVOT and might be subjected to pressure trauma by the valve frame. The relative area of the region of interest that experiences contact pressure is defined as contact pressure index and a contact pressure index of >14% was defined as the cut-off point for conduction abnormalities. (11)

A subsequent computational fluid dynamic simulation of the blood flow in diastole is computed to predict the risk of PVL. A blood flow of >16mL/s was defined as a cut-off point for moderate PVL. (12)

### Outcomes and definitions

The primary objective of the study was to evaluate to what extent computer simulations in challenging aortic anatomies may affect TAVR sizing and implantation strategies and identify patients at risk for high degree AV blocks or more than trace PVL. The follow-up period was 30 days. PVL-assessment was performed by TTE.

### Statistical analysis

Distribution of continuous variables were tested for normality with the Shapiro-Wilk test. Continuous variables were reported as mean ± standard deviation or median (25^TH^ −75^TH^ percentile) and analyzed with a student’s T-test, ANOVA, Mann Whitney U-or Kruskal-Wallis-test as appropriate. Categorical variables were reported as percentage and compared with Chi-Square or Fishers Exact test. The best-fitted simulation, based on the implantation depth, for each patient was used to evaluate correlation with PVL and new pacemakers. Receiver-operating characteristic (ROC)-curves were generated to find the optimal cut-off values for >trace PVL based on the computer model PVL-measurements and for new PPI post-TAVR based on the computer model contact pressure index. (Youden index criteria). A 2-sided P-value <0.05 was considered statistically significant. All statistics were performed with SPSS software version 28.0 (SPSS, Chicago IL, United States).

## Results

### Study population

MSCT studies of 83 patients were transmitted for FEops HEARTguide™ computer simulations. Quality was insufficient for computer simulations in 6 cases. Therefore, the study cohort consisted of 77 patients undergoing a TAVR-procedure between October 2020 and April 2022. Baseline characteristics are depicted in Table 1. Median age was 79.9 years (25^th^-75^th^ percentile 74.2-83.8), 42% was male, median BMI was 27.0 kg/m^2^ (25^th^-75^th^ percentile 22.8-34.0) and median Surgeon’s Predicted Risk of Mortality (STS-PROM) was 2.8% (25^th^-75^th^ percentile 1.8 −4.1) with clinical frailty in 35%. MSCT-analysis revealed a mean annulus area of 443mm^3^ (±91.3) and a severely calcified aortic valve in 74% of the patients. The mean Agatston score was 4405 ± 978 in male patients and 2824 ± 1368 in female patients. The challenging anatomy was a bicuspid valve in 17 patients (22%), a small annulus in 13 (17%) patients, and a severely calcified tricuspid valve in 47 patients (61%).

**Table 1:**
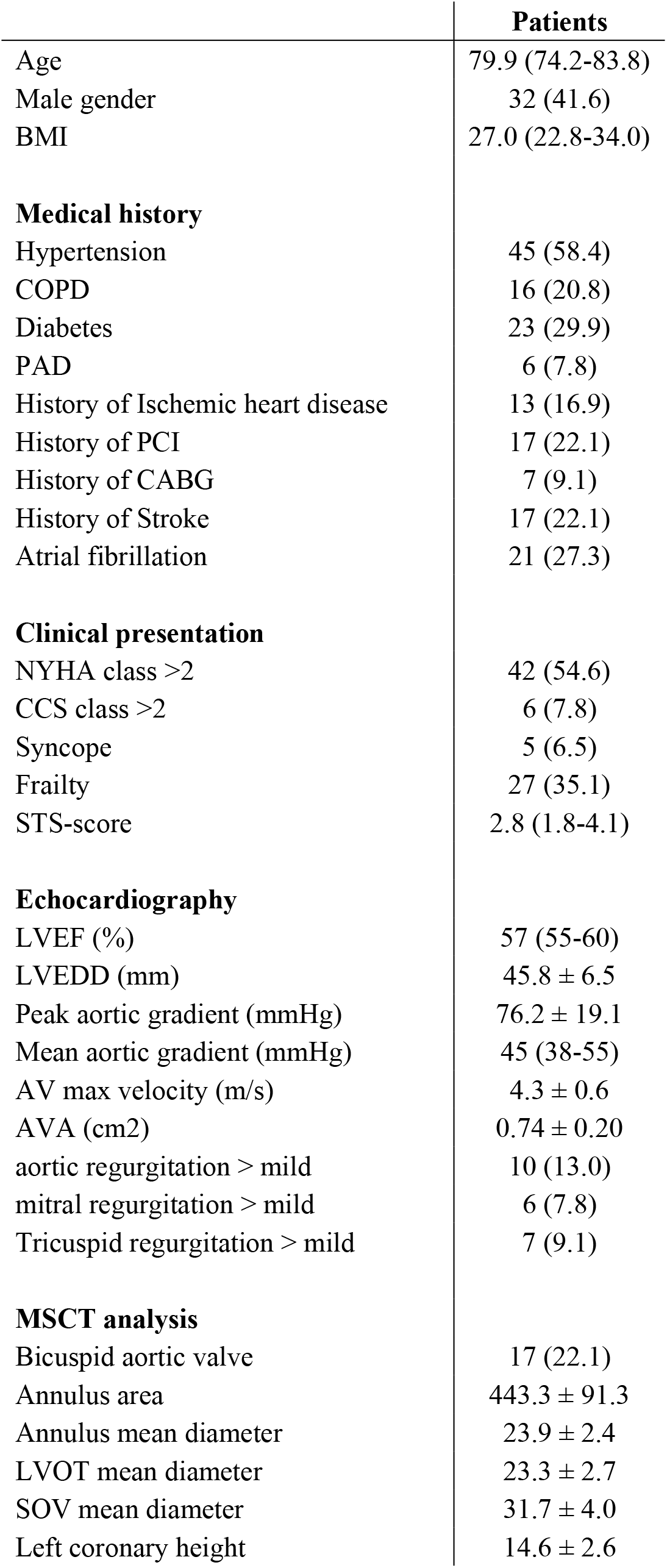
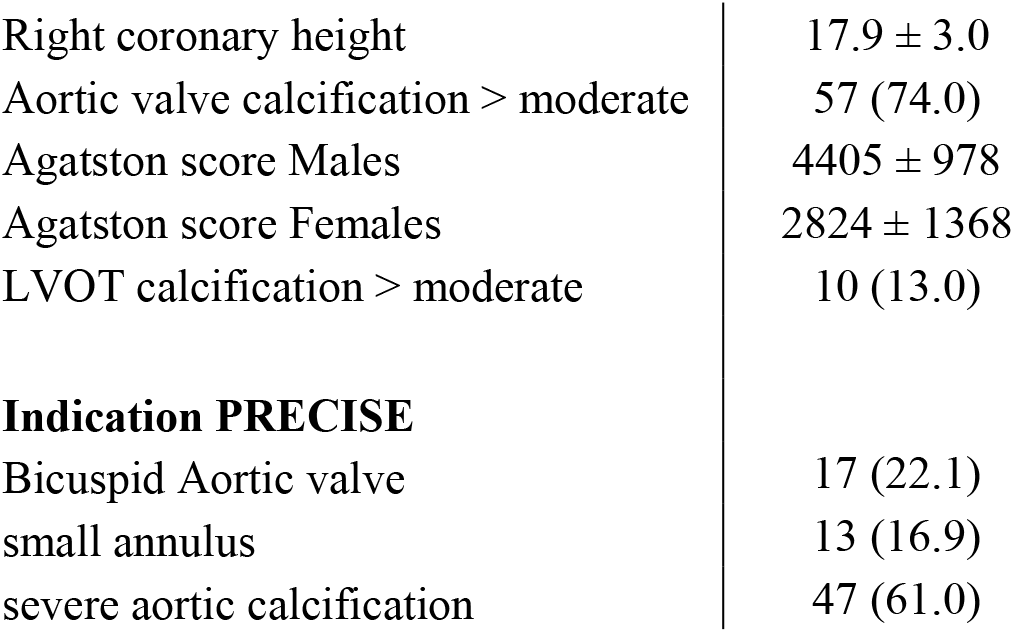
baseline characteristics. BMI = Body Mass index, COPD = Chronic Obstructive Pulmonary Disease, PAD = Peripheral Artery Disease, PCI = Percutaneous Coronary Intervention, CABG = Coronary Artery Bypass Graft, NYHA = New York Heart Association, CCS = Canadian Cardiovascular Society, STS = Society of Thoracic Surgeons, LVEF = Left Ventricular Ejection Fraction, LVEDD = Left Ventricular End Diastolic Dimensions, AV = Aortic Valve, AVA = Aortic Valve Area, LVOT = Left Ventricular Outflow tract, SOV = Sinus of Valsalva

|  | **Patients** |
| --- | --- |
| Age | 79.9 (74.2-83.8) |
| Male gender | 32 (41.6) |
| BMI | 27.0 (22.8-34.0) |
| **Medical history** |  |
| Hypertension | 45 (58.4) |
| COPD | 16 (20.8) |
| Diabetes | 23 (29.9) |
| PAD | 6 (7.8) |
| History of Ischemic heart disease | 13 (16.9) |
| History of PCI | 17 (22.1) |
| History of CABG | 7 (9.1) |
| History of Stroke | 17 (22.1) |
| Atrial fibrillation | 21 (27.3) |
| **Clinical presentation** |  |
| NYHA class >2 | 42 (54.6) |
| CCS class >2 | 6 (7.8) |
| Syncope | 5 (6.5) |
| Frailty | 27 (35.1) |
| STS-score | 2.8 (1.8-4.1) |
| **Echocardiography** |  |
| LVEF (%) | 57 (55-60) |
| LVEDD (mm) | 45.8 ± 6.5 |
| Peak aortic gradient (mmHg) | 76.2 ± 19.1 |
| Mean aortic gradient (mmHg) | 45 (38-55) |
| AV max velocity (m/s) | 4.3 ± 0.6 |
| AVA (cm<sup>2</sup>) | 0.74 ± 0.20 |
| aortic regurgitation > mild | 10 (13.0) |
| mitral regurgitation > mild | 6 (7.8) |
| Tricuspid regurgitation > mild | 7 (9.1) |
| **MSCT analysis** |  |
| Bicuspid aortic valve | 17 (22.1) |
| Annulus area | 443.3 ± 91.3 |
| Annulus mean diameter | 23.9 ± 2.4 |
| LVOT mean diameter | 23.3 ± 2.7 |
| SOV mean diameter | 31.7 ± 4.0 |
| Left coronary height | 14.6 ± 2.6 |
| Right coronary height | 17.9 ± 3.0 |
| Aortic valve calcification > moderate | 57 (74.0) |
| Agatston score Males | 4405 ± 978 |
| Agatston score Females | 2824 ± 1368 |
| LVOT calcification > moderate | 10 (13.0) |
| **Indication PRECISE** |  |
| Bicuspid Aortic valve | 17 (22.1) |
| small annulus | 13 (16.9) |
| severe aortic calcification | 47 (61.0) |

### Procedure and 30-day outcomes

Pre-procedural planning changed after computer simulations in 35% of cases (change in valve size selection (12%) or target implantation height (23%)). Procedural characteristics are shown in Table 2. Predilatation was performed in 62% and postdilatation in 27%. Evolut size was 23mm in 5%, 26mm in 31%, 29mm in 46% and 34mm in 18%. Valve migration occurred in 2 (3%) patients, a second valve was necessary in 3 (4%) patients and conversion to surgery in 1 (1%) patient.

**Table 2:**
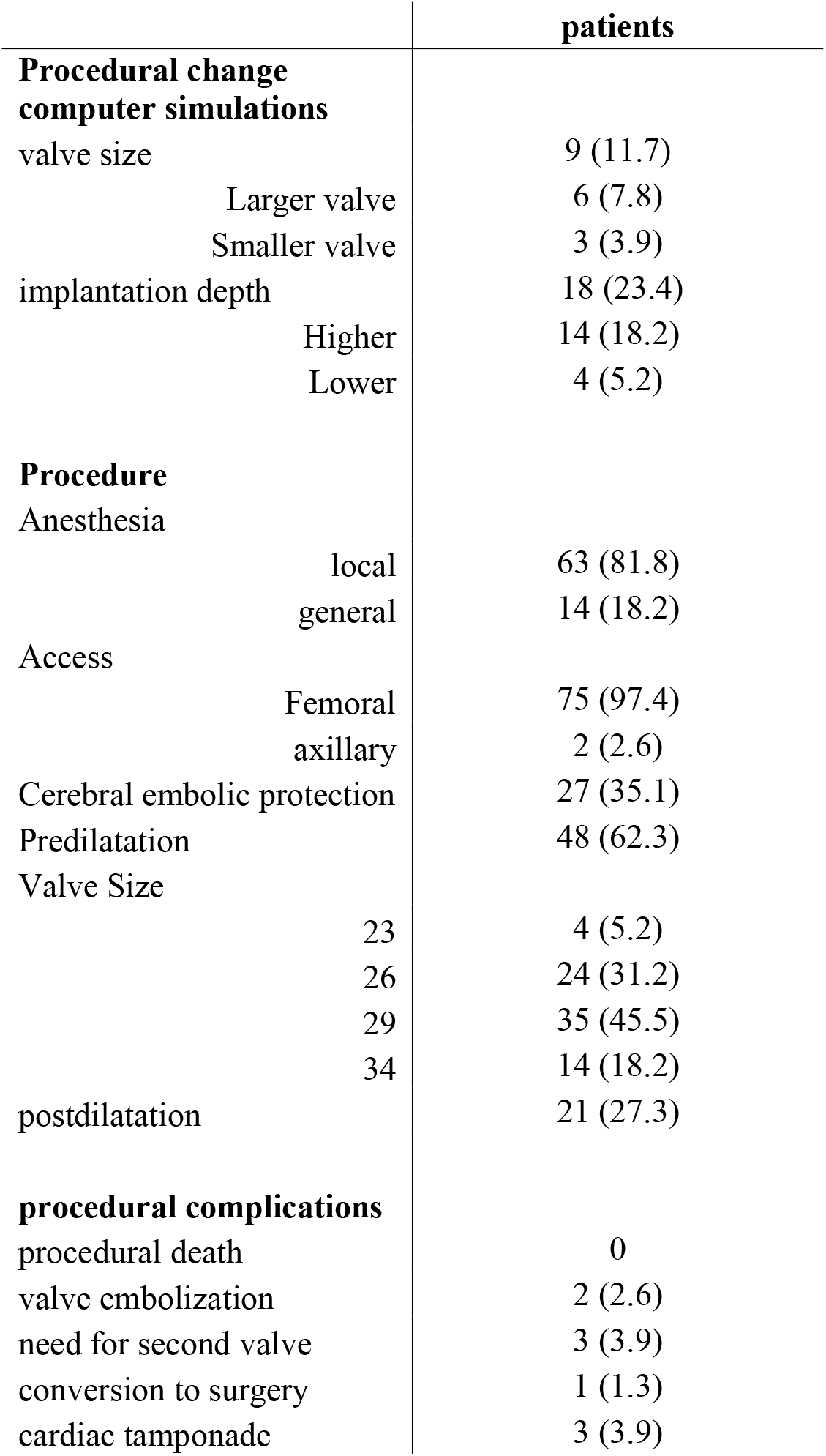
procedural characteristics. characteristics of the TAVR procedure.

The 30-day outcomes are displayed in Table 3. New LBBB occurred in 14% and a permanent pacemaker was implanted in 10 patients (13%) (total AV block in 9 patients and a brady-tachy syndrome in 1). The implantation depth relative to NCC, as measured by angiography, was 5.6±3.8 for the patients without a new PPI versus vs. 5.7±4.0 for the patients who received a new PPI (p=0.71). Echocardiography post-TAVR showed none-trace PVL in 71.1% of the patients, mild PVL in 22% and moderate PVL in 5 patients (6.5%).

**Table 3:** 30-day Outcomes: AVB = atrioventricular block, LBBB = Left Bundle Branch Block, RBBB = Right Bundle Branch Block, LVEF = Left Ventricular Ejection Fraction, LVEDD = Left Ventricular End Diastolic Dimensions, AV = Aortic Valve, AVA = Aortic Valve Area, PVL = Paravalvular leakage

|  | Patients |
| --- | --- |
| Death | 1 (1.3) |
| Myocardial Infarction | 0 |
| Disabling Stroke | 1 (1.3) |
| Acute Kidney Injury | 1 (1.3) |
| Major bleeding | 3 (3.9) |
| Major vascular complication | 6 (7.8) |
| Pacemaker implantation | 10 (13.0) |
| **ECG** |  |
| Rhythm |  |
| sinus rhythm | 52 (70.3) |
| Atrial fibrillation | 14 (18.2) |
| Paced | 8 (10.4) |
| AV block |  |
| new 1st degree AVB | 9 (11.7) |
| Bundle Branch Block |  |
| new LBBB | 11 (14.2) |
| new RBBB | 4 (5.2) |
| **Echocardiography** |  |
| LVEF (%) | 59 (55-60) |
| LVEDD (mm) | 47.2 ± 5.6 |
| Peak aortic gradient (mmHg) | 15 (10-21) |
| Mean aortic gradient (mmHg) | 8 (5-11) |
| AV max velocity (m/s) | 1.9 (1.6-2.3) |
| AVA (cm<sup>2</sup>) | 1.7 (1.6-2.4) |
| PVL |  |
| None-trace | 55 (71.4) |
| Mild | 17 (22.1) |
| moderate | 5 (6.5) |
| mitral regurgitation > mild | 5 (6.5) |
| Tricuspid regurgitation > mild | 6 (7.8) |

### Computer simulations

The computer simulations that more closely matched the procedure in terms of valve size and implantation depth were compared with the relevant clinical events. The contact pressure index was significantly higher in patients with a new PPI, compared to those without (16.0% (25^th^-75^th^ percentile 12.0-21.0) vs. 3.5% (25^th^-75^th^ percentile 0-11.3), p<0.01) (Figure 1). A cut-off value of 11.5% correlated well with PPI (AUC 0.83 (95% confidence interval (CI) 0.72-0.94), sensitivity 86%, specificity 76%). Two patients (4.3%) with a contact pressure index ≤14% and 5 patients (31.3%) with a contact pressure >14% received a new PPI (p<0.01).

**Figure 1:**
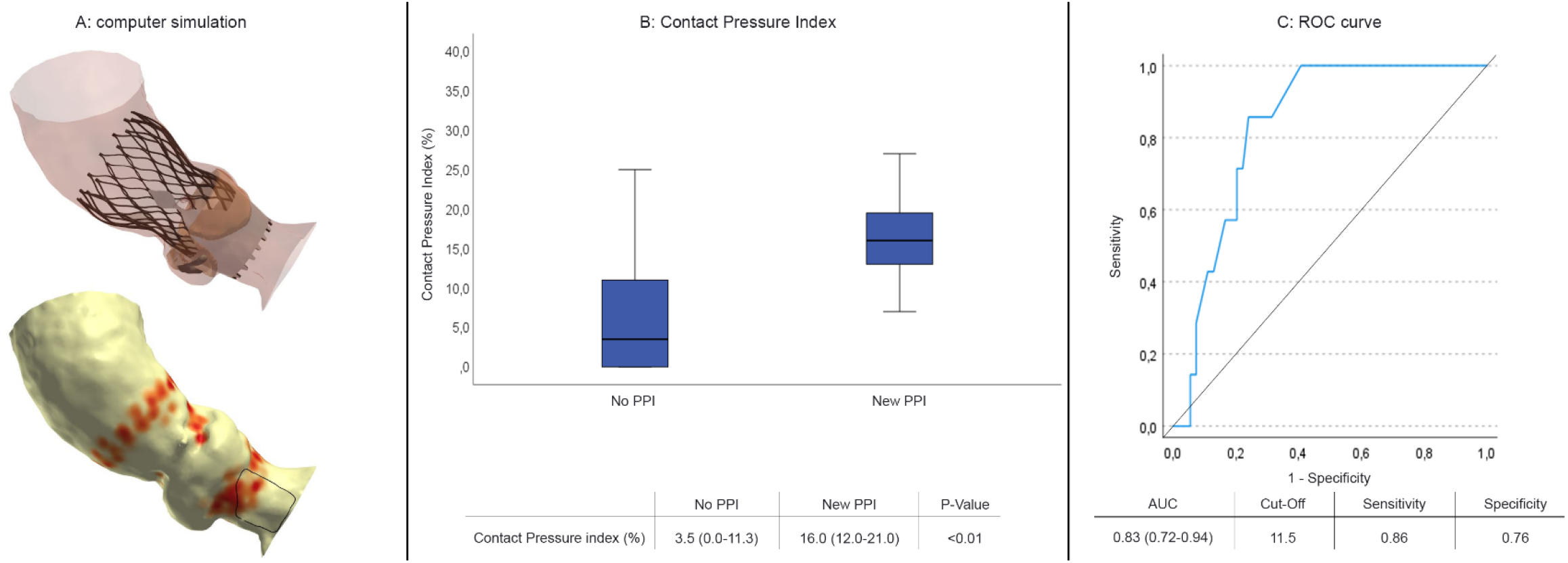
PPI-prediction. Prediction of permanent pacemaker implantations. Panel A: visualization of the FEops HEARTguide simulation: (top) prediction of the frame deformation after THV deployment, (bottom) visualization of the contact pressure (red spots) exerted on the aorta. The contact pressure index is defined as the relative area of contact within the region of interest (in black) and used as an indication for risk of conduction abnormalities. Panel B: the median contact pressure index, Panel C: the ROC curve to predict new PPI. PPI = permanent pacemaker implantation, AUC = Area under the Curve

Figure 2 illustrates the PVL-analysis. The predicted PVL was 5.7mL/s (25^th^-75^th^ percentile 1.3-11.1) in patients with none-trace PVL, 12.7 (25^th^-75^th^ percentile 5.5-19.1) in mild PVL and 17.7 (25^th^-75^th^ percentile 3.6-19.4) in moderate PVL. A PVL cutoff of 12.2 mL/s helped discriminating patients with > trace PVL (AUC 0.69 (95% CI 0.55-0.82), sensitivity 59%, specificity 79%). Eleven patients (19.3%) with a PVL simulation ≤16mL/s and 11 patients with a PVL >16 mL/s (61.1%) had a PVL > trace (p<0.01) (supplemental table 1).

**Figure 2:**
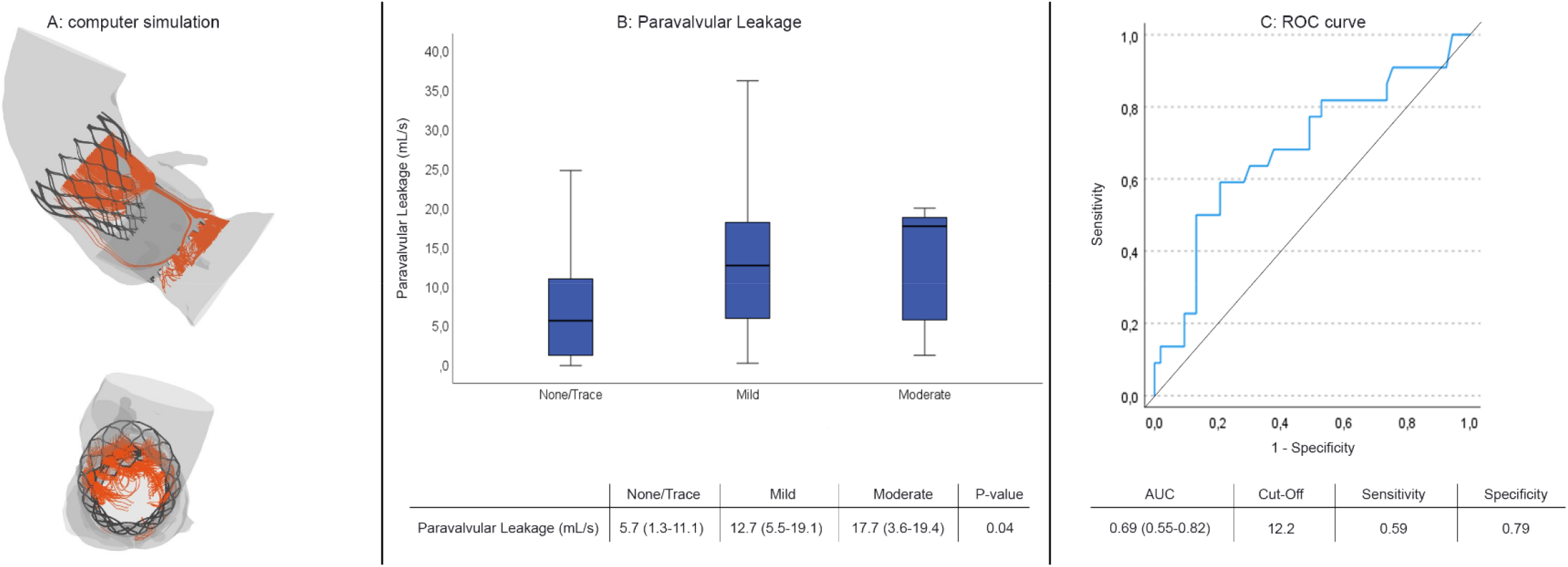
PVL-prediction. prediction of paravalvular leakage: Panel A: Visualization of the FEops HEARTguide simulation for prediction of PVL: PVL leak is shown as orange streamlines from a front view (top) and a top view (bottom). Panel B: the median predicted PVL per PVL-grade, Panel C: the ROC curve to predict new >trace PVL. PVL = Paravalvular leakage, AUC = Area under the Curve

## Discussion

PRECISE-TAVR is the first multicenter, prospective study to evaluate the added value of computer simulations to pre-procedural planning in patients with a challenging anatomy receiving an Evolut Pro-valve. The main findings are that computer simulations with FEops HEARTguide™ 1) changed the pre-procedural planning in 35% of the patients, 2) well predicted the risk for PVL and 3) identified the risk for new PPI post-TAVR.

Pre-procedural planning is increasingly important for optimal sizing and to identify anatomical risk factors for adverse events.(3) Optimal valve size selection leads to a proper THV fit in the native anatomy ensuring adequate hemodynamic valve performance, proper sealing with no PVL and avoiding excessive contact pressure in the LVOT that may result in conduction disorders.

Conventional MSCT imaging tools do not consider device-host interactions. Indeed, aortic root shape (elliptical vs circular, vertical vs. horizontal, long vs. short LVOT) and calcifications (location, amount/volume) may affect how a THV is deployed and seated in situ. Arguably, device-host interactions may be reinforced in more complex anatomical phenotypes such as bicuspid aortic valves, heavily calcified tricuspid valves, and small anatomies. Severe calcifications of the aortic valve lead to frame eccentricity post-TAVR which enhances the severity of PVL. (8) Also, high calcium burden, especially in the left coronary cusp (LCC) is a risk factor for PPI.(9) TAVR in patients with BAV is associated with multiple procedural adverse events, including a higher moderate-severe PVL-rate and new PPI, especially in patients with a calcified raphe.(10,15) In small anatomies, self-expandable valves are hemodynamically superior to balloon-expandable valves, however the >trace PVL rate remains high between 49% and 75%.(16,17)

Our study demonstrates that computer simulations of these device-host interactions may complement procedural planning. Local heart teams changed their strategy in a third of the patients to help mitigate PVL and conduction disorders in complex anatomies. Computer simulations prompted operators to change size and to aim for a higher THV implantation in 12% and 23% of the cases respectively. The 13% PPI rate should be seen in the context of complex anatomies where reported PPI rates vary between 14 and 26%.(9,15,18)

In our study, FEops HEARTguide™ simulations predicted > trace PVL fairly well. The new defined PVL cutoff of 12.2 mL/s is lower than the 16mL PVL cutoff reported in the FEops HEARTguide™ validation study. However, this 12.2mL cutoff in PRECISE-TAVR discriminated < trace from > trace PVL whereas the 16mL threshold in the validation study was used to predict > mild PVL (sensitivity 0.72 and specificity 0.78). Interestingly, in our study a PVL > 16 mL/s was associated with > trace PVL in 61% of cases vs. only 19% when PVL flow < 16mL/s. A specific threshold for > trace PVL seems relevant as mild PVL may also be associated with longer hospitalizations and mortality.(19) Furthermore, we identified a step-wise increased in computer-derived PVL flow as post-procedural PVL severity increased, consistent with recent findings.(20) The incidence of > trace PVL in our study is similar to what has been reported in the post-market FORWARD Pro study with 41% > trace PVL and 2% moderate-severe PVL in a population that was not selected for its complex anatomy but rather aimed to reflect every day practice.

In PRECISE-TAVR, a contact pressure index of 11.5 % was a good predictor for PPI post-TAVR (sensitivity 0.86, specificity 0.76) and compares with the 14% threshold in the original validation study for any conduction abnormalities (PPI or new LBBB) (sensitivity 0.95, specificity 0.54).(11) In our study a contact pressure > 14% resulted in a PPI rate of 31% vs. 4% when contact pressure remained ≤14%. Contact pressure will increase with deeper THV implantation. Mean depth of implantation (DOI) was 5.7±4.0 for patients with PPI vs. 5.6±3.8mm for patients with no PPI, showing that implantation depth alone is not a good predictor for PPI post-TAVR. The PPI rate in PRECISE-TAVR was 13% and compares favorably with PPI rates in post-market registries (FORWARD 18%; FORWARD Pro 19%), a RCT in low-risk patients with PPI (17%) and comparable to a propensity-matched analysis using new implantation techniques(12%).(7,21-23) New implantation techniques such as the double S curve and the cusp overlap technique were not systematically applied in PRECISE-TAVR. Intuitively, a cusp overlap technique should result in higher DOI with corresponding lower contact pressure and potentially lower PPI. The length of the membranous septum also correlates with PPI. In particular a short MS (< 3mm) is associated with a higher PPI because there is a higher likelihood of contact interference between the THV frame and the His Bundle.(24)

FEops HEARTguide™ not only considers MS length but also accounts for the interaction of the THV with the surrounding anatomical structures resulting in contact pressure on the region of the conduction system. This contact pressure may correlate better with PPI than MS length per se.

## Limitations

PRECISE-TAVR is a prospective observational study that only considered the Evolut Pro platform in patients with predefined complex anatomies. There was no independent screening committee, and all patients were selected by the respective local heart teams. The decision to perform additional manoeuvres to correct PVL or to proceed with PPI was at the discretion of the treating physician. Of note, total AV block was identified in 90% of patients with PPI. Echocardiograms after TAVR were evaluated by local imagers and not by an independent echocardiography Core Laboratory. Finally, computer simulations predict what ideally would happen for a specific THV size and implantation depth in a specific anatomy. Operators may not always manage to implant the THV in the exact same location as suggested by the simulation. Also, a one-shot implantation may have a different effect than multiple repositioning attempts that may result in more device LVOT interactions and trauma.

## Conclusion

Feops HEARTguide™ simulations may provide enhanced insights in the risk for PVL or PPI after TAVR with a self-expanding supra-annular THV in complex anatomies.

## Data Availability

Data is available by request to the corresponding author

## Abbreviations

BAV: Bicuspid Aortic Valve
ECG: Electrocardiogram
LVOT: Left Ventricular Outflow Tract
MSCT: Multislice Computed Tomography
PPI: Permanent pacemaker Implantation
PVL: Paravalvular Leakage
TAVR: Transcatheter Aortic Valve Replacement
TTE: Transthoracacic Echocardiography

## Specific requests

### What is known

Conventional pre-procedural planning in patient undergoing TAVR includes a contrast-enhanced multi-slice computed tomography for obtaining detailed information on aortic valve morphology, dimensions, calcium burden, membranous septum length, LVOT and transcatheter heart valve sizing. However, device-host interactions remains difficult to predict, especially in patients with a challenging anatomy, leading to a higher risk of permanent pacemaker implantation and paravalvular leakage.

### What the study adds

The PRECISE-TAVR trial showed that the use computer simulations leads to a significant change in pre-procedural planning, including valve size and implantation height. When comparing the implantation depth post-TAVR with the computer simulations, FEops HEARTguide™ simulations accurately predicts which patients are at risk for PPI or >trace PVL.

### Sources of Funding

PRECISE-TAVR is part of a project that has received funding from the European Union’s Horizon 2020 research and innovation program under grant agreement No. 945698.

### Disclosures

- Thijmen W Hokken: none
- Henrik Wienemann: none
- James Dargan: none
- Dirk-Jan van Ginkel: none
- Cameron Dowling reports grants from Medtronic, outside the submitted work.
- Axel Unbehaun: none
- Johan Bosmans works as clinical proctor Medtronic CoreValve
- Andreas Bader-Wolfe: none
- Robert Gooley reports personal fees from Boston Scientific, outside the submitted work.
- Martin Swaans reports proctoring fees for training/educational services to the Department of Cardiology from Boston Scientific, Abbott Vascular, Edwards Lifesciences, Cardiac dimensions, Philips Healthcare and Bioventrix inc.
- Stephen J. Brecker reports Consultancy fees from : Medtronic & Aortic Innovations
- Matti Adam reports grants and personal fees from Medtronic, and personal fees from JenaValve, Edwards Lifesciences, and Boston Scientific
- Nicolas M. Van Mieghem has received research support from Abbott, Boston Scientific, Edwards Lifesciences, Medtronic, PulseCath, Daiichi Sankyo, Pie Medical, Materialise and consultancy fees from Abbott, Anteris, Boston Scientific, Medtronic, Abiomed, PulseCath, Daiichi Sankyo, Teleflex

---

**Supplemental table 1:** table to show the differences in PPI and PVL in the validated cut-off points. PPI = permanent pacemaker implantation, CPI = contact pressure index, PVL = paravalvular leakage

|  | **Validated cut-off points** |  |
| --- | --- | --- |
|  | **CPI ≤14%** | **CPI >14%** |
| **New PPI** | 2 (4.3) | 5 (31.3) |
|  | **PVL ≤16mL/s** | **PVL >16mL/s** |
| **> Trace PVL** | 11 (19.3) | 11 (61.1) |

